# Early Identification of High-Risk Individuals for Mortality after Lung Transplantation: A Retrospective Cohort Study with Topological Transformers

**DOI:** 10.1101/2025.10.01.25337124

**Authors:** Alexy Tran-Dinh, Enora Atchade, Sébastien Tanaka, Brice Lortat-Jacob, Yves Castier, Hervé Mal, Jonathan Messika, Pierre Mordant, Philippe Montravers, Ian Morilla

**Affiliations:** Université Paris Cité, AP-HP, Hôpital Bichat Claude Bernard, Département d’anesthésie-Réanimation, INSERM, Paris, France; Université Paris Cité, LVTS, Inserm U1148, F-75018 Paris, France; UMR 1188, INSERM, Université de la Réunion, Saint-Denis de la Réunion, France; Université Paris Cité, AP-HP, Hôpital Bichat Claude Bernard, Département de chirurgie thoracique et vasculaire, Paris, France; Université Paris Cité, Inserm U1152, F-75018 Paris, France; Université Paris Cité, AP-HP, Hôpital Bichat Claude Bernard, Pneumologie B et Transplantation Pulmonaire, Paris, France; Paris Transplant Group, Paris, France; Université Sorbonne Paris Nord, LAGA, CNRS, UMR 7539, Laboratoire d’excellence Inflamex, F-93430, Villetaneuse, France; Instituto de Hortofruticultura Subtropical y Mediterránea “La Mayora” (IHSM-UMA-CSIC), Genetics Area, Faculty of Sciences, University of Malaga, Málaga, Spain

**Keywords:** Lung transplantation, Mortality risk, Topological transformers, Machine learning, Predictive modeling

## Abstract

**Background and Objective:** Lung transplantation remains the only definitive treatment for patients with end-stage respiratory failure; however, it is burdened by a substantial risk of post-operative mortality. Current risk stratification methods, such as the Lung Transplant Risk Index, offer limited predictive performance and interpretability. This study introduces a novel predictive model based on topological transformers to assess mortality risk following lung transplantation. The objective is to improve predictive accuracy by capturing complex temporal patterns in clinical data while ensuring model interpretability to inform clinical decisions.

**Methods:** A retrospective cohort study was conducted using clinical data from lung transplant recipients. The model integrates both static and time-dependent clinical variables through a transformer-based architecture that incorporates topological features derived from patients’ temporal trajectories. Model performance was compared to established methods using a held-out test set. The evaluation metrics included accuracy, sensitivity, specificity, and the area under the receiver operating characteristic curve. Model interpretability was assessed using Shapley Additive explanations to identify and rank the most influential predictors of mortality.

**Results:** The proposed model demonstrated superior predictive performance compared to the Lung Transplant Risk Index and other benchmark models. On the test dataset, it achieved an accuracy of 87.4%, sensitivity of 84.1%, and specificity of 89.6%. The model consistently outperformed existing approaches across different subgroups, including age, underlying disease, and transplant type. Shapley-based interpretability analysis revealed that dynamic variables such as early post-operative oxygenation trends, immunosuppressive load, and inflammatory markers were among the most critical contributors to mortality risk.

**Conclusions:** The integration of topological features within a transformer-based framework significantly enhances the prediction of post-transplant mortality risk. By offering both improved predictive power and model transparency, this approach supports more precise and personalised risk stratification in lung transplantation. These findings highlight the potential of topological transformers as a valuable tool in the broader context of precision medicine and clinical decision support.

## Introduction

Lung transplantation remains a critical intervention for patients with end-stage lung disease, yet post-transplantation mortality within the first year remains a significant challenge, with rates ranging from 10% to 20%^1^. Existing predictive models, such as LTRI and the Charlson Comorbidity Index, exhibit limitations due to their static nature and lack of incorporation of temporal clinical adaptative changes. Previous research has explored risk prediction models for lung transplantation, primarily focusing on static clinical scores. These scores included data as age, lung function, diagnosis, and comorbidities^2,3,4,5,6^. Yusen^2^ introduced LTRI, which considers preoperative factors but lacks real-time adaptability. Similarly, Rajkomar^7^ demonstrated ML applications in medical predictions, yet their models do not incorporate TDA-based transformations. Recent studies have applied ML to transplant-related outcomes, but none have leveraged topological transformers to model patient trajectories dynamically. In general, the existing predictive models, exhibit limitations due to their static nature and lack of incorporation of temporal clinical adaptative changes. This study introduces a predictive framework that combines topological data analysis (TDA) with machine learning (ML) to capture dynamic patient trajectories. Unlike conventional risk assessment models, this approach encodes both preoperative and postoperative variables into a structured topological representation, enhancing predictive accuracy and clinical applicability^7^. This study then advances in the development of a dynamic risk assessment model using topological transformers, outperforming existing scoring systems such as Cox regression^8^ or the ex vivo lung perfusion (EVLP)^9,10^. As well as, it Integrates multi-scale topological descriptors into ML classifiers, providing a more robust feature representation. It also validates the model’s effectiveness through a retrospective cohort study and comparison with standard clinical indices. To, finally, enhances the model interpretability using SHapley Additive exPlanations (SHAP) values, ensuring clinical relevance and applicability.

Our goal was to significantly ameliorate the precision of predicting one-year mortality risk after lung transplantation. To achieve this, we conducted a retrospective analysis of a cohort of over 251 patients who underwent lung transplantation at Bichat Claude’s Bernard hospital in Paris over the last five years. Yet, we took advantage of a recently novel approach^11^ implementing a partial encoding of the topological transformers described in it. Using these insights^12,13,14^, we developed a predictive model for one-year mortality risk after lung transplantation that achieved significantly higher precision than existing models.

Our results demonstrated that our approach outperformed existing models and achieved higher accuracy in predicting the risk of mortality after lung transplantation. The interpretability of our models, as revealed by SHAP values, indicates that our approach provides actionable insights for clinical decision-making. The large sample size and extensive clinical data collected in our study provide a robust foundation for future research in this area.

## Methods

### Study Design, Setting, and Participant Cohort

A retrospective cohort of 252 lung transplant recipients was analyzed, with 189 survivors and 63 non-survivors. Specifically, between 2015 and 2020, pre-, intra- and post-operative characteristics of all consecutive patients undergoing lung transplantation at Bichat Hospital (Paris, France) were prospectively collected, including age, sex, body mass index, comorbidities (ischemic heart disease, diabetes mellitus, preoperative pulmonary arterial pressure, etiology of lung disease, mismatch for cytomegalovirus serology, pre-operative ECMO, lung transplantation in high emergency procedure, single or double lung transplantation, cold ischemic time, thoracic epidural analgesia, intraoperative ECMO, transfusions, SOFA *(Sequential Organ Failure Assessment)* score and SAPS 2 *(Simplified Acute Physiology Score 2)* at postoperative admission in intensive care unit (ICU), number of days on norepinephrine, mechanical ventilation and ECMO, length of ICU stay, the onset of grade-3 primary graft dysfunction, acute renal failure, bacteremia, pneumonia, pleural empyema, bronchial stenosis, bronchial anastomosis dehiscence, antibody-mediated rejection and cellular rejection and the need for tracheotomy.

### Data Preprocessing and Nonlinear Dimensionality Reduction

We used heat diffusion to capture the underlying geometry of our high-dimensional data, with the goal of preserving the intrinsic structure of the data.

To this end, we performed a modified PHATE (Potential of Heat-diffusion for Affinity-based Transition Embedding) nonlinear dimensionality reduction and data projection^15^. Briefly, we constructed a Markov chain from the high-dimensional data, where each data point represents a state in the chain, and the transition probabilities between states were determined by the pairwise similarities between the data points. We adjust the graph construction parameters to capture the appropriate level of connectivity between patients. Heat diffusion is then used to smooth out the transition probabilities over a range of time scales, which helps to reveal the underlying geometry of the data.

Once the Markov chain has been constructed, this algorithm uses a variant of spectral clustering to embed the data into a lower-dimensional space (see Fig.1b top). At this point, we experiment with different embedding dimensions to find the optimal balance between preserving the underlying geometry of the data and reducing its dimensionality, allowing for meaningful visualizations and downstream analysis (see Fig. 1b down).

**Figure 1.**
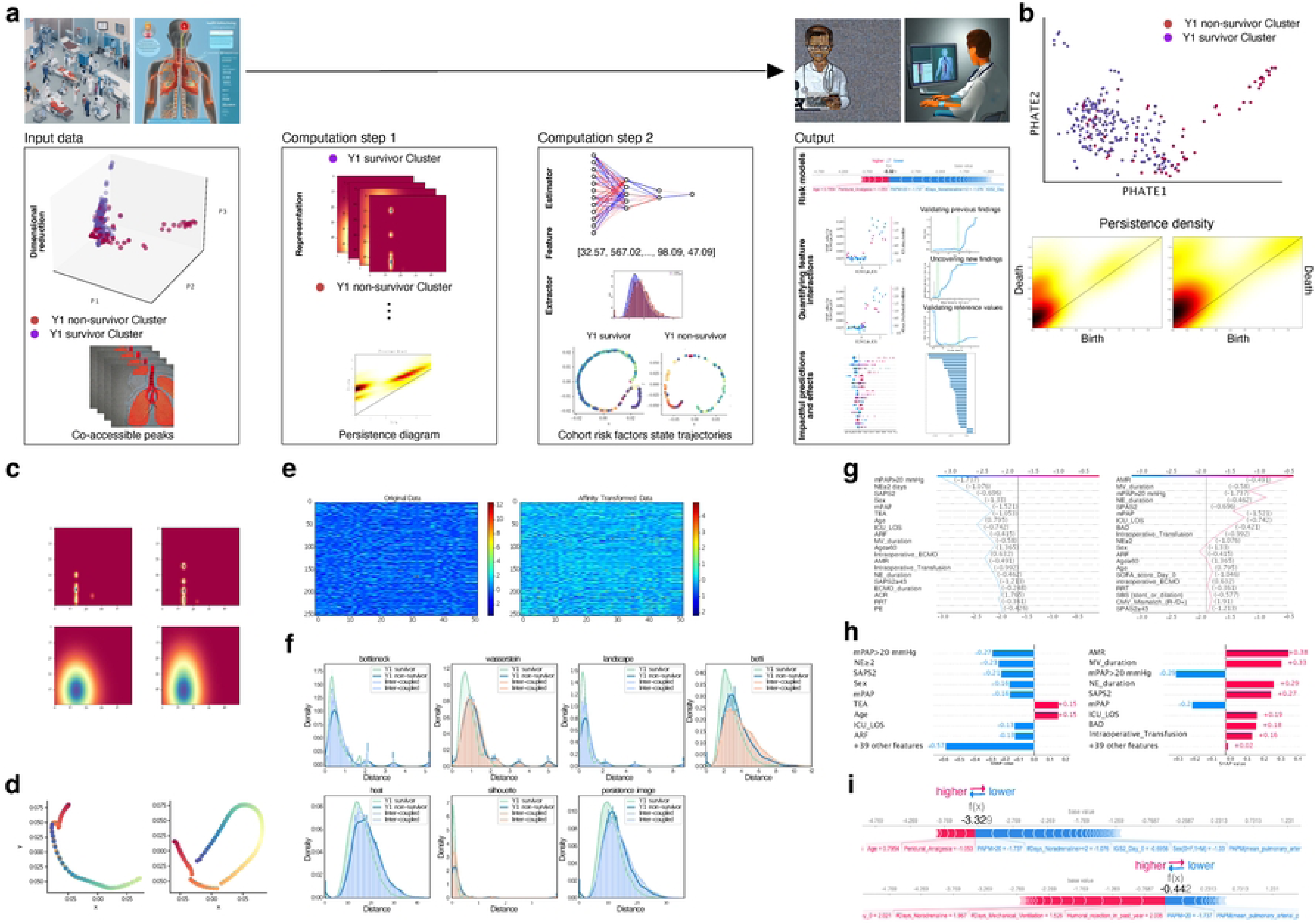
Overview of Topological Transformers and Their Application in Identifying High-Risk Mortality in Year One After Lung Transplantation. In panel a, we started with inpatient data and visualized the data records for each co-accessible peak per lung recipient over time. We then used a non-linear manifold to represent this data in a way that is accessible to medical users. Next, in computation step 1, we identified important clinical latent variables and encoded them using a single vector of topological extractors that maintain the structure of the data. We used this topological vector as an estimator in a machine learning predictor to predict mortality risk in computation step 2. Additionally, we were able to use the topological extractors to track cohort risk factor trajectories dynamically. Finally, we predicted the impact of each variable and quantified their interaction effects on the risk model of Y1 mortality. Based on the interpretability of the obtained learning model, we assigned a risk score to each patient, providing a valuable tool for clinicians in managing the care of lung transplant recipients. **b**, shows the construction of a low-dimensional embedding of data patient while clusters samples by a diffusion process that fits well the spotted branching trajectories of our data. We transform clinical variables into diagrams of persistence visualized by densities and localize the most suitable candidates to be homology generators of dimension 0, 1, and 2 in data patient. In panel **c**, the homology generators are used as tiled regions where to extract important clinical latent variables. We show a frequent situation of image persistence extractor at different pixel resolutions for survivor and non-survivor patients on the left and right, respectively. **d**, shows the trajectory inference between survivor and non-survivor clusters. **e**, visualizes transformed data patients by the picked topological transformer using the matrix of affinity calculated after applying the non-linear learning manifold technique. A smooth version of the data can be retrieved after affinity application, as shown in panel **f**, which also demonstrates the effect of each topological transformer intra and inter clusters. Frequent decision-making depending on the local interpretability of the risk model is demonstrated in panel **g**, with survivor and non-survivor patients on the left and right, respectively. Panel **h** shows the same with regard to the local effect of clinical variables. Finally, panel **i** demonstrates personalized early detection of the risk of mortality in Y1 according to patient local and global clinical variable interactions. The top and bottom individuals represent survivor and non-survivor patients, respectively. Red indicates a high feature value and blue indicates a low feature value. In the context of a binary classification problem (survivor/non-survivor), red bars may indicate that a certain feature increases the probability of surviving, while blue bars may indicate that the feature decreases the probability of surviving. Overall, the topological transformers provide an innovative and effective approach to identifying high-risk mortality in lung transplant recipients, with the potential to improve clinical decision-making and patient outcomes.

### Topological Feature Extraction using Persistent Homology

This section describes the steps taken to perform TDA using the giotto-tda Python package^16^ in tailored Python scripts to create persistence diagrams and persistence images^17,18,19^. We then used the persistence images to vectorise the data and build a multilayer perceptron (MLP) model in scikit-learn library for classification^20^.

Topological data analysis was used to analyse the topological features of the data. TDA is a mathematical framework that enables the extraction of topological features from complex data sets. In this study, we applied persistent homology, a technique within TDA, to identify invariant topological features in our data. To do this, we used the software package, Giotto-TDA version 0.5.1, which implements the computation of persistent homology. The data was first preprocessed to ensure that it was suitable for TDA analysis. We then constructed a simplicial complex from the data, which was used to compute the persistent homology. The persistent homology diagrams and densities were generated using GUDHI version 3.7.0^21^ and were used to identify topological features, such as holes and voids, in the data (see Fig. 2c-d). Finally, statistical analyses were performed on the identified topological features to test their significance. Specifically, to compare the relative importance of different topological features within a persistence diagram, we quantified the magnitude of a topological feature, such as a peak or a hole, in a persistent homology diagram using the number of points, the persistence entropy, and amplitude feature extractors (see Fig. 3). The amplitude measures the vertical distance between the maximum and minimum points on the persistence diagram corresponding to a given feature representation, namely: bottleneck, Wasserstein, landscape, betti curve, heat, silhouette, and persistence image^19^. This feature extractor is particularly useful in distinguishing between peaks or holes with similar persistence (i.e., lifespan) but different amplitudes. The number of points provides information about the overall size and density of the dataset, while the persistence entropy extractor provides information about the complexity and diversity of the topological features in the dataset.

**Figure 2.**
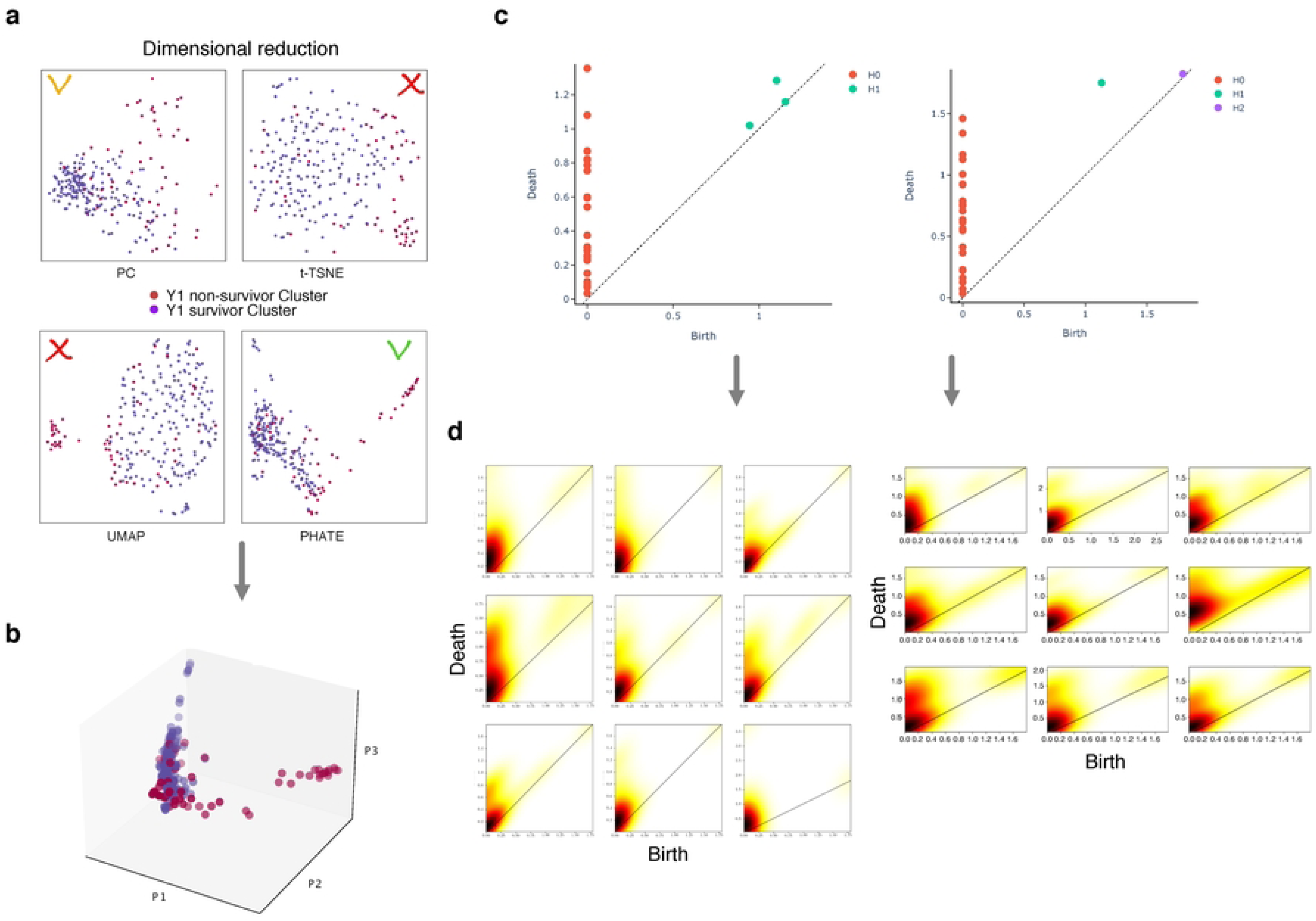
Overview of Lower-Dimensional Visualizations and Feature Contribution to the Prediction for Lung Transplantation Patients. **a**, lower-dimensional visualisation of lung transplant patients using t-SNE, UMAP, PCA, and adapted PHATE techniques. Each dot represents a patient, and the colors indicate the level of acceptance of the lung transplant, with red indicating not accepted, orange fairly accepted, green accepted and recommended. **b**, 3D rotated visualisation of inpatients using the same techniques as that accepted in a. The plot provides a clearer separation between the groups of patients with different transplant acceptance levels. **c**, feature contribution to the prediction of lung transplant patient outcomes across pseudo temporal scales. The plot shows two typical examples, one for a survivor (left-hand side) and one for a non-survivor (right-hand side). Each feature’s contribution is displayed as a persistence diagram (see methods), with red, green, and purple indicating a high contribution at different topological resolution. **d**, feature contribution to the prediction of lung transplant patient outcomes across spatial scales. The plot shows the same two typical examples as in **c**, but the x-axis represents the spatial position of the individual topological feature. The plots reveal the spatial distribution of these features and their importance in predicting the risk of mortality after transplantation.

**Figure 3.**
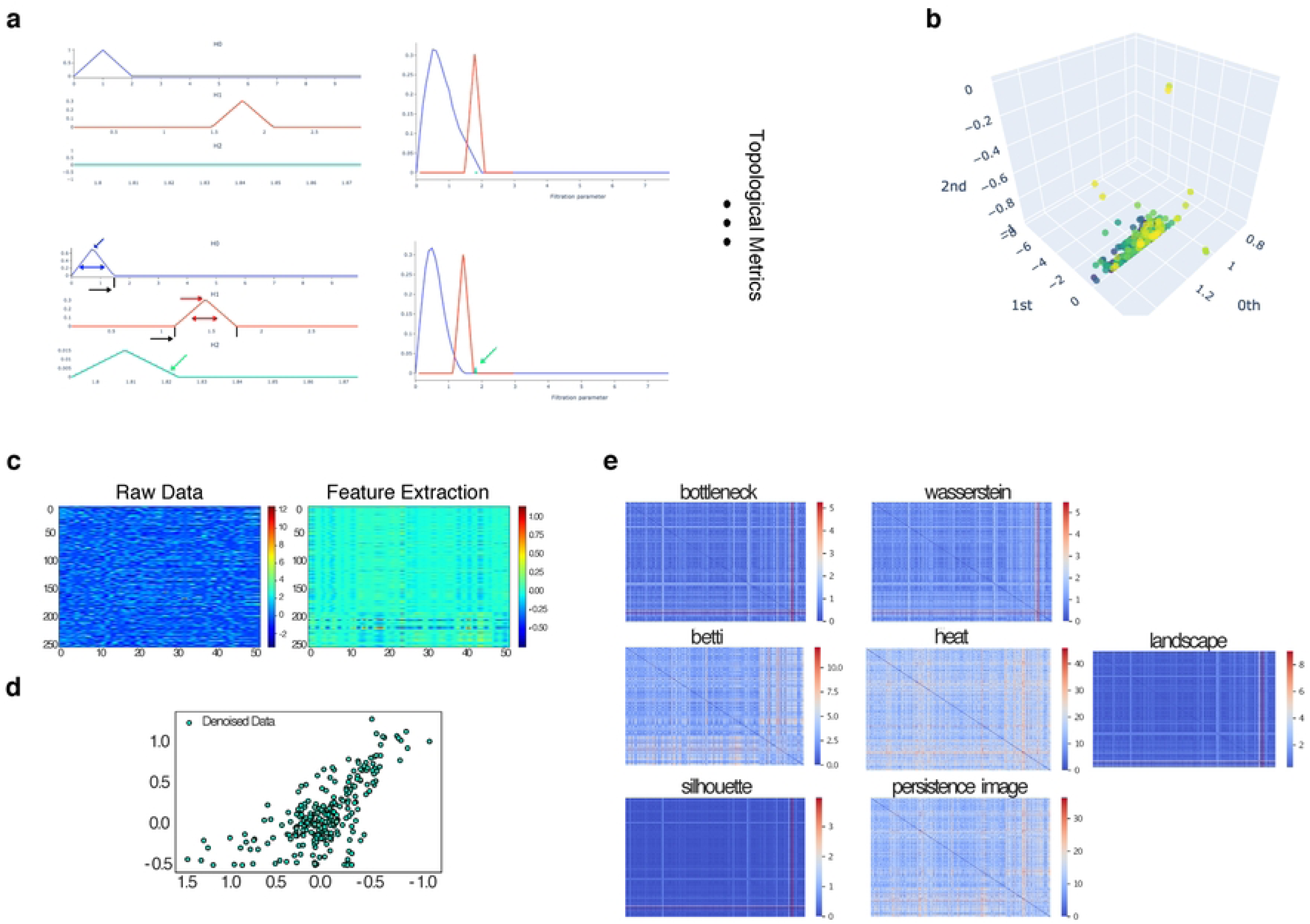
Topological Metrics and Extractors. **a**, left: topological metrics throughout the entire filtration process. The highlighted arrows in the bottom-left indicate persistence increases with a later and more dilated trend in non-survivor individuals. Furthermore, homology generators of dimension 2 are mostly absent in survivor cases, as shown in the top-left. Right: Silhouettes show a more dilated and later appearance with no clustered structure of data in the dimension 2 generators. These trends are consistent throughout the rest of the metrics, such as heat-kernel and persistence image. **b**, entropy of the persistence diagram provides information about the level of variability in the data and how it evolves over time. **c-d**, heatmaps of the transformed patient-by-variable matrix using different topological extractors. The color bar ranges from 0 to 1, and the heatmap is smooth, with colors ranging from −0.5 to 0. **e**, heatmaps of the pairwise distance matrices between patients using different topological extractors. Each heatmap represents a different extractor and allows for visual comparison between them.

### Machine Learning Model Development and Training

We employed persistence images as representations of the persistence diagram to enable efficient topological data analysis. Thus, we fine-tuned the persistence images to a 100 x 100-pixel resolution with a spread of 1.0. By visualizing the data’s topological features in this way, we were able to conduct a more thorough analysis of the inpatients’ data. These representations were then vectorised and used as input to train a scikit-learn Multi-Layer Perceptron (MLP) model^21^. This vectorisation technique involves transforming the persistence images and other representations into a fixed-length feature vector, which can be used as input to a machine learning model. Our model supported as well other topological feature representations as persistence landscapes or Betti curves and provides a range of options for tuning the parameters of the learning model algorithms (see Fig. 3a-b). It was generated a single feature vector composed by 3 + 3 + (7 x 3) = 27 topological features, by concatenating 9 features per homology dimension (see Fig. 4a). The vectorisation approach was based on the work of Brunner et al. (2020)^22^, while the use of giotto-tda and MLP models for topological data analysis was supported by prior work such as Carrière et al. (2020)^16^ and Fletcher et al. (2020)^23^.

**Figure 4.**
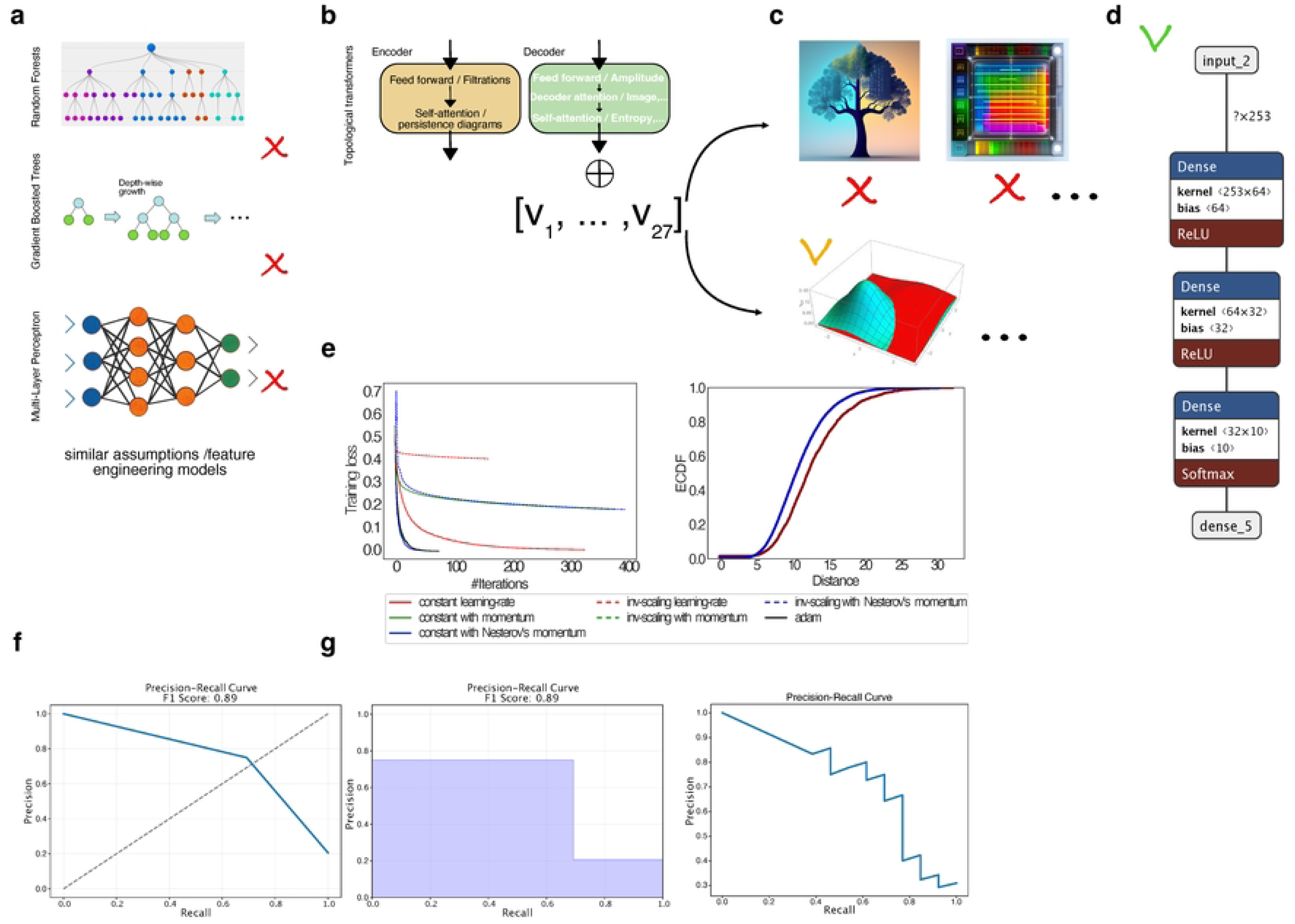
Performance evaluation of the predictive model. **a**, classification results using three different feature engineering models (Random Forests, Gradient Boosted Trees, and Multi-Layer Perceptron) without topological transformers. b) Integration of extracted features using topological transformers. c) Comparison of different approaches and their evaluations, including Multi-Layer Perceptron, which outperformed all other examined classification algorithms. d) Table 1 showing the evaluation metrics of the different approaches, including accuracy, precision, recall, and F1-score. e) Left: the best result for learning on the patient dataset using different training algorithms. Right: an ECDF plot of the distance values for two groups of samples, Y1 survivors, and Y1 non-survivors. f) Evaluation curves re-calibrated based on the previously described distance, including a subplot showing the performance of the machine learning model in terms of precision and recall, as well as the corresponding F1-score, and a subplot showing the precision-recall curve displayed differently using a step function. g) The trade-off between precision and recall for a binary classification model.

**Table 1.** Alternative learning models (LMs) that were compared to our MLP, along with their corresponding performances and topological features. The table shows that all cases with an accuracy above 80% still perform significantly worse than our MLP. For instance, the Gaussian Process result was computed with default optimizer and Adaboost*, while Quadratic Discriminant Analysis yielded some collinear variables**. Despite these attempts, our MLP model still outperformed them all.

| <u>LMs</u> | **Random Forest** | **Gaussian Process** | **Radial Basis Function** | **Gaussian Naive Bayes** | **Decision Tree** | **Quadratic Discriminant Analysis** | **KNeighbours** |
| --- | --- | --- | --- | --- | --- | --- | --- |
| <u>Accuracy</u> | 0.81 | 0.79/0.66* | 0.79 | 0.8253 | 0.76 | 0.8095 ** | 0.8253 |

To classify the lung transplantation patients based on their mortality risk, we used a multilayer perceptron (MLP) model (see Fig. 4d). By means of one input layer, one or more hidden layers, and one output layer this type of artificial neural network can learn complex nonlinear relationships in the data^21^. The input layer receives the input data, which is then processed by the hidden layers to learn important features of the data, and finally produces the output through the output layer.

Each neuron in the hidden layers and output layer applies an activation function to introduce non-linearity and capture complex patterns in data. MLP learns the weights of the connections between neurons through backpropagation, which adjusts the weights to minimize the difference between predicted and actual output. During our exploration of different classification algorithms, we evaluated several different approaches, such as Gaussian Process, Radial Basis Function (RBF), K-Neighbors, Gaussian Naive Bayes, Decision Tree, and Quadratic Discriminant Analysis^21^. Finally, our analysis revealed that MLP outperformed all the other classification algorithms that we examined. However, MLP is known to be susceptible to overfitting, especially when the number of hidden layers or neurons is relatively large compared to the amount of training data available. To address this issue, regularization techniques such as dropout and weight decay are commonly employed. Nevertheless, we propose a novel approach that utilizes topological invariants to prevent overfitting, which avoids the need for these conventional regularization techniques.

Topological invariants are mathematical properties that are invariant under continuous transformations of a topological space. In the context of machine learning, they can be used to represent the underlying structure of the data, such as its shape or connectivity, and can be used to build a more robust and interpretable model.

Unlike traditional regularization techniques, which add constraints or penalties to the model to prevent overfitting, topological invariants use the inherent structure of the data to ensure that the model generalizes well. Specifically, they provide a way to measure the complexity of the model and its ability to capture the essential features of the data, while avoiding the risk of overfitting.

By incorporating topological invariants into the design of the model, we can achieve a better balance between its capacity to capture the complexity of the data and its ability to generalize to new, unseen data points. This approach can help to prevent overfitting, without the need for traditional regularization techniques, which can be computationally expensive and difficult to tune.

### Dynamic Trajectory Analysis using Topological Transformers

In the context of studying within-subject lung transplantation state trajectories (see Fig.5), persistence images could be used to analyze changes in transplant over time and how they relate to the particular to health outcomes that subjects are experiencing, such as lung function, survival, and incidence of rejection. We discretized persistence images of lung transplantation states for each time step, which creates a matrix that represents the topological activity of the transplant states at each time point.

**Figure 5.**
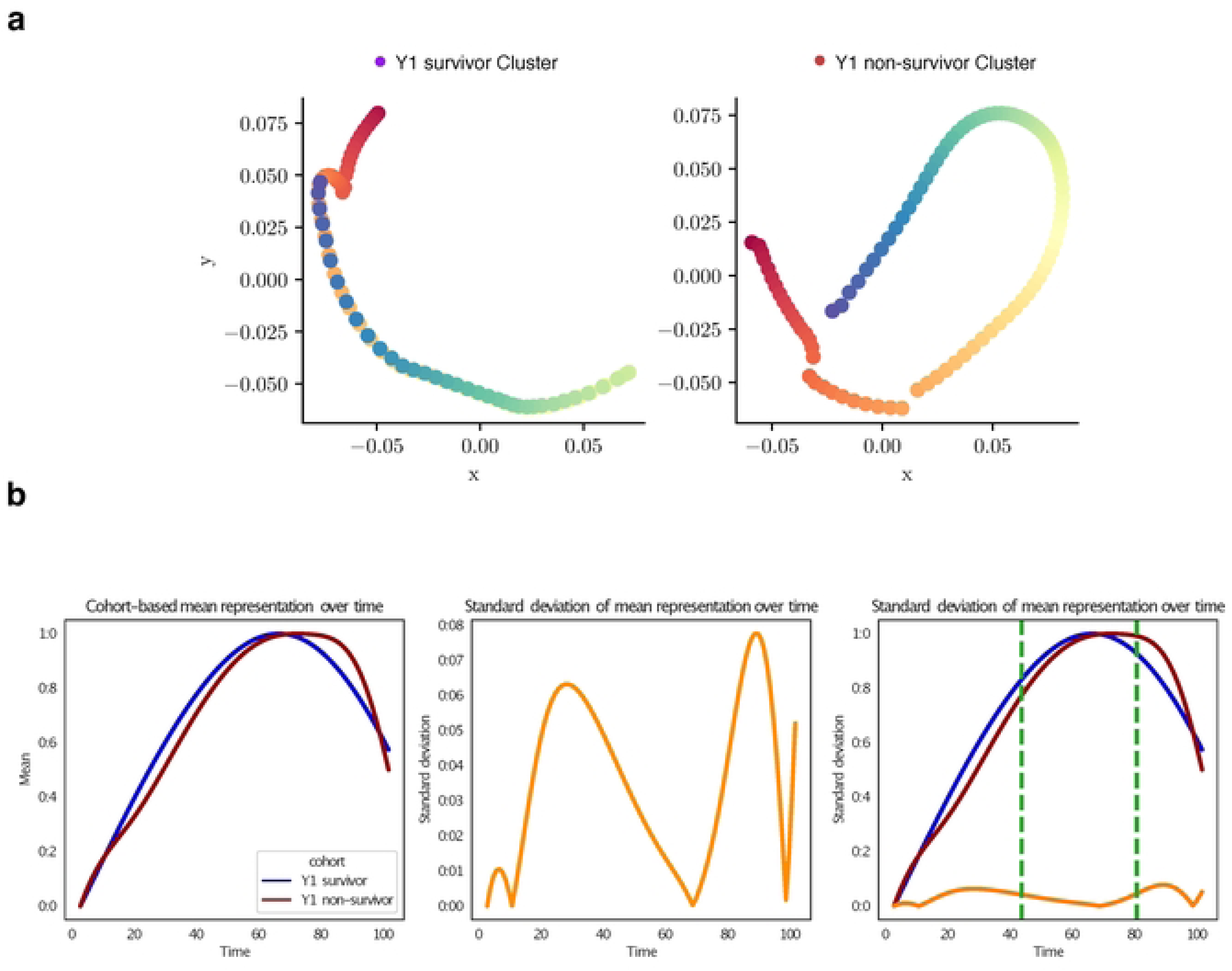
Trajectories of ECMO variables in survivor and non-survivor individuals aged 15-71. **a**, trajectory plots of ECMO variables in survivor and non-survivor individuals. The left panel shows the trajectories for survivors, with a linear pattern indicating a simpler course of ECMO therapy. The right panel shows trajectories for non-survivors, with greater variability and complexity in the trajectory. **b**, variability analysis between survivor and non-survivor individuals. Subplot 1 shows survival probability curves for survivor and non-survivor individuals, with the survivor curve surpassing the non-survivor curve at the point of their maxima. Subplot 2 shows a curve of standard deviation as a function of cluster labels, with three normal mixture distributions plotted on the curve. Subplot 3 shows the relationship between standard deviation, mean, and time in the dataset, with vertical lines at time points indicate important events or changes in the dataset.

These matrices could then be used to calculate the sample mean of each participant cohort, resulting in a matrix whose rows represent the average topological activity of the lung transplantation states for participants in the respective cohort. Taking the Euclidean distance between persistence images as a proxy for their actual topological dissimilarity, we calculated pairwise distances between rows of each matrix and embed them using an adapted phate algorithm for time-varying data^24^.

By transforming persistence diagrams into images, you could create a dataset of time-varying images that is used to train our machine learning model to predict health outcomes or identify patterns of lung transplantation that are associated with certain outcomes.

Overall, the use of persistence images could provide a powerful and flexible tool for analysing complex time-varying data and identifying patterns and trends that might be missed by other methods. However, it’s important to carefully consider the appropriate choices for weight functions and probability distributions to ensure that the resulting images are both informative and unbiased.

Finally, all this resulted in a 2D trajectory of the lung transplantation states over time (see Fig.5a), where the state is measured using topological extractors. By analyzing these trajectories, we could identified patterns and trends in the transplantation states that are associated with the outcomes that subjects are suffering from. This information could be used to better understand the underlying mechanisms that contribute to the patient’s outcome, and to develop more targeted interventions or treatments.

We quantify the variability across cohorts, our algorithm reads in a dataset and a CSV file containing cluster labels for each observation in the whole cohort. It then computes the mean representation for each cluster over time and normalises the mean representation between 0 and 1. The code then visualises the cohort-based mean representation over time, the standard deviation of the mean representation over time, and the cohort-based mean representation separately over time again. Finally, we visualise the indices where certain salient features in the dataset have the highest effect.

### Model Interpretability and Risk Score Derivation

Interpretable machine learning models were developed to identify key features associated with mortality risk in our study population. We used a Multilayer Perceptron (MLP) architecture, which is a type of neural network that has been shown to be effective in predicting medical outcomes^25^. We fine-tuned the model using topological invariants, which are mathematical representations of the spatial relationships between data points^19^.

To make the MLP models interpretable, we used the SHAP Python library^14^. We employed several approaches provided by SHAP to further investigate the effect of individual features on mortality risk.

First, we created a heatmap of SHAP values, which visualizes the contribution of each feature for each individual in the dataset. This helped us to identify features that were consistently important across the entire dataset.

Next, to further investigate the effect of individual features on mortality risk, we generated individual-level SHAP summary plots. These plots illustrate the contribution of each feature to the predicted mortality risk for a given individual. By examining these plots, we were able to identify key features that were consistently associated with increased mortality risk, such as age, ECMO in ICU, and comorbidities^14^.

The SHAP dependence plots (see Figure 6d-e) are a powerful visualization technique used to understand the interactions between the features in a machine learning model. Specifically, these plots show the relationship between a selected feature and the top three features that may interact with it in the model. To generate these plots, the SHAP library is used to calculate the contribution of each feature to the model’s prediction. Then, the interaction strengths between the selected feature and the other features in the dataset are estimated, and the top three features with the strongest interactions are chosen for plotting.

**Figure 6.**
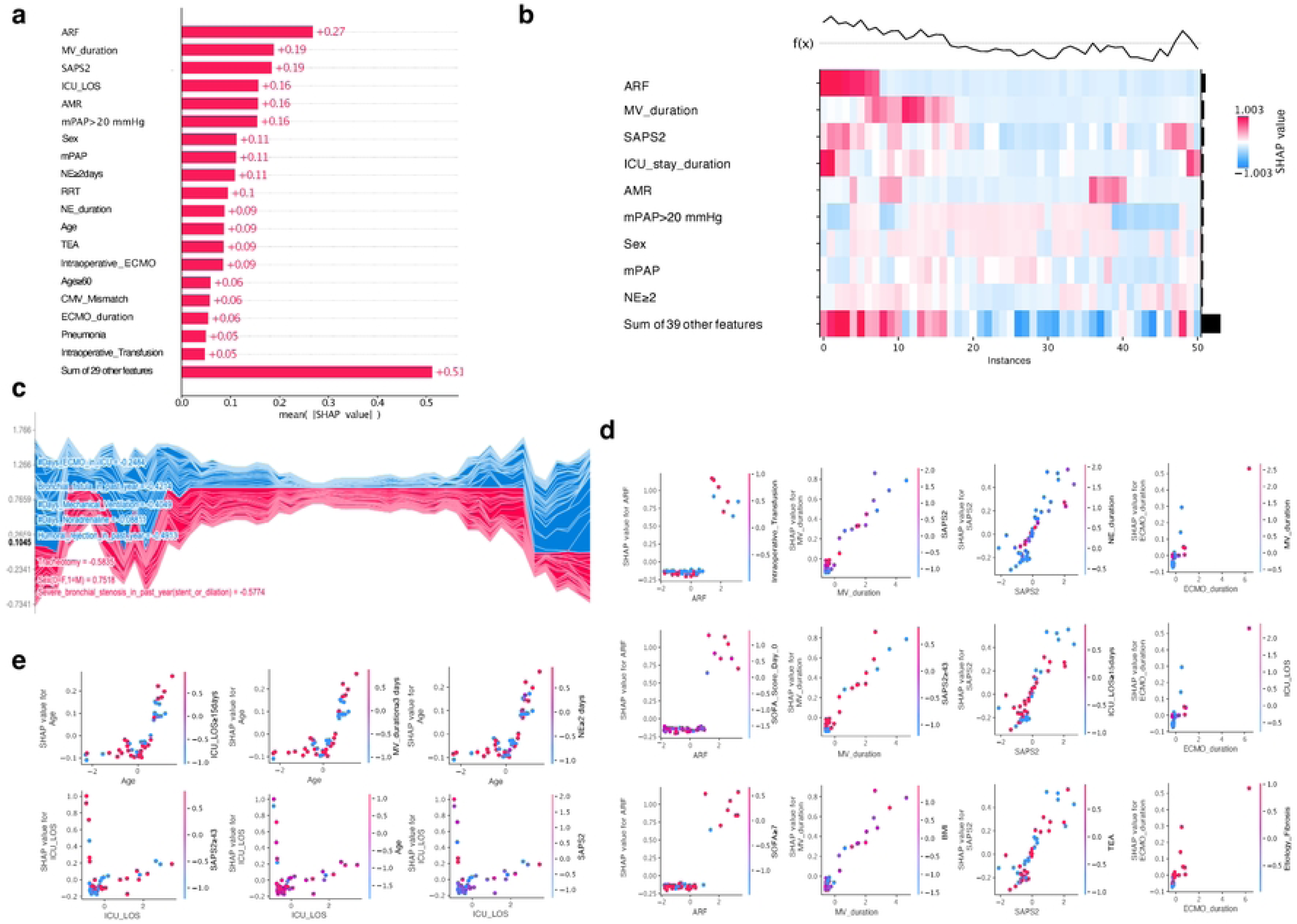
Feature contribution to prediction. **a**, SHAP bar plot showing the impact of each feature on the model output, in terms of its contribution to the prediction. Red bars indicate features that increase the prediction, while blue bars indicate features that decrease it. The top 5 features with the largest impact on the predictions are presented, in order of importance. **b**, heatmap of the top 10 features individually, while grouping the remaining 39. Positive values are typically shown in shades of red, while negative values are represented in shades of blue. **c**, SHAP force plot that describes the feature contributions to the model prediction for each individual instance in the cohort. **d**, SHAP dependence plot illustrating the local feature interactions for the top 3 most impactful variables at only one different interacting feature value each time. **e**, SHAP dependence plot for ICU stay duration, showing a complex relationship with interacting features.

Each SHAP dependence plot shows how the selected feature impacts the model’s prediction for different values of the interacting feature. The interaction effect is represented by the color of the points in the plot, while the vertical position of the points represents the SHAP value of the selected feature. The plots are arranged in a 3×3 grid, with each row representing a different interacting feature and each column representing a different range of values for that feature.

For example, if the top three interacting features are “age”, “heart rate”, and “temperature”, then the first row of the plot would show the interaction between the selected feature and “age” in the three columns, with each column representing a different age range. The second row would show the interaction between the selected feature and “heart rate” in the three columns, with each column representing a different heart rate range. Finally, the third row would show the interaction between the selected feature and “temperature” in the three columns, with each column representing a different temperature range.

Furthermore, we utilised SHAP values to visualise and comprehend how the model arrived at its predictions for individual patients. We examined the contribution of each feature to the predicted mortality risk for a single patient and probed the model about the reasoning behind its decision based on the values of each feature for that particular patient.

In addition, we investigated the relationship between individual features and mortality risk while controlling for the effects of other features. This allowed us to better understand how individual features contribute to the overall prediction of mortality risk.

Finally, we calculated the Cohen’s distance (*d*) effect size to measure the strength of association between each feature and mortality risk^26^. This helped us to prioritize which features to focus on in our analysis (see Figs. 6-7).

**Figure 7.**
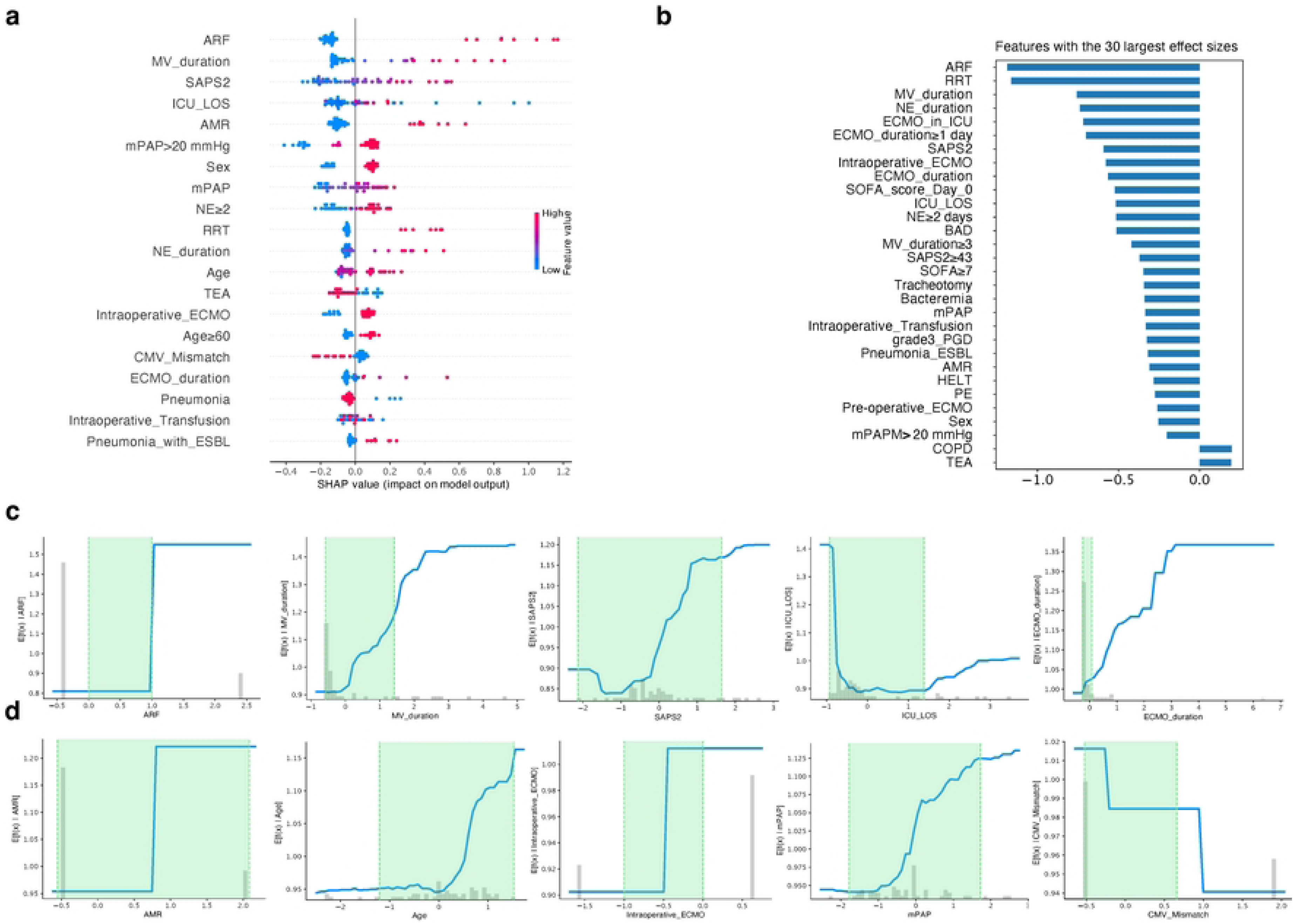
Contribution to model output. **a**, impact of different features on the model output. The red values indicate positive feature values that have a positive impact on the model output, while the remaining values are more centered around 0 and have a less significant impact. The top 5 features with the highest SHAP values are ARF, MV duration, SAPS2 Day 0, ICU_LOS, and AMR. **b**, comparison of swarm plot, shap bar plot, and Cohen’s distance effect bar plot to identify important features for the model output. The consistent important features are identified, and their effect on the model output is explored. **c**, partial dependence plots for ARF, MV duration, SAPS2 Day 0, ICU_LOS, and ECMO duration. The shaded regions represent the level of confidence in the model’s predictions, with a wider shaded region indicating greater variability in the model’s predictions. **d**, partial dependence plots for Age and the recently discovered important COPD variable, along with three other feature variables. The subplots suggest the presence of non-linear relationships between some features and the model output.

Overall, the use of interpretable machine learning models, specifically the MLP architecture fine-tuned by topological invariants and the SHAP Python library with these additional approaches, allowed us to identify and understand the key features associated with mortality risk in our study population.

### Statistical Analysis and Performance Evaluation

From the MLP interpretability analyses it is created a score model based on the feature importance values of our machine learning model. The SHAP values provided a measure of how much each input variable contributed to the overall output of the model. We then visualised those values to identify the features that had the highest values and therefore the greatest impact on the output of the model.

To create the score model, we assigned a weight to each feature based on its SHAP value, which was calculated as the absolute value of the SHAP value divided by the sum of the absolute values of all SHAP values. We then computed the score for each patient in our dataset by summing the weighted values for each of the most important features (see Fig. 8).

**Figure 8.**
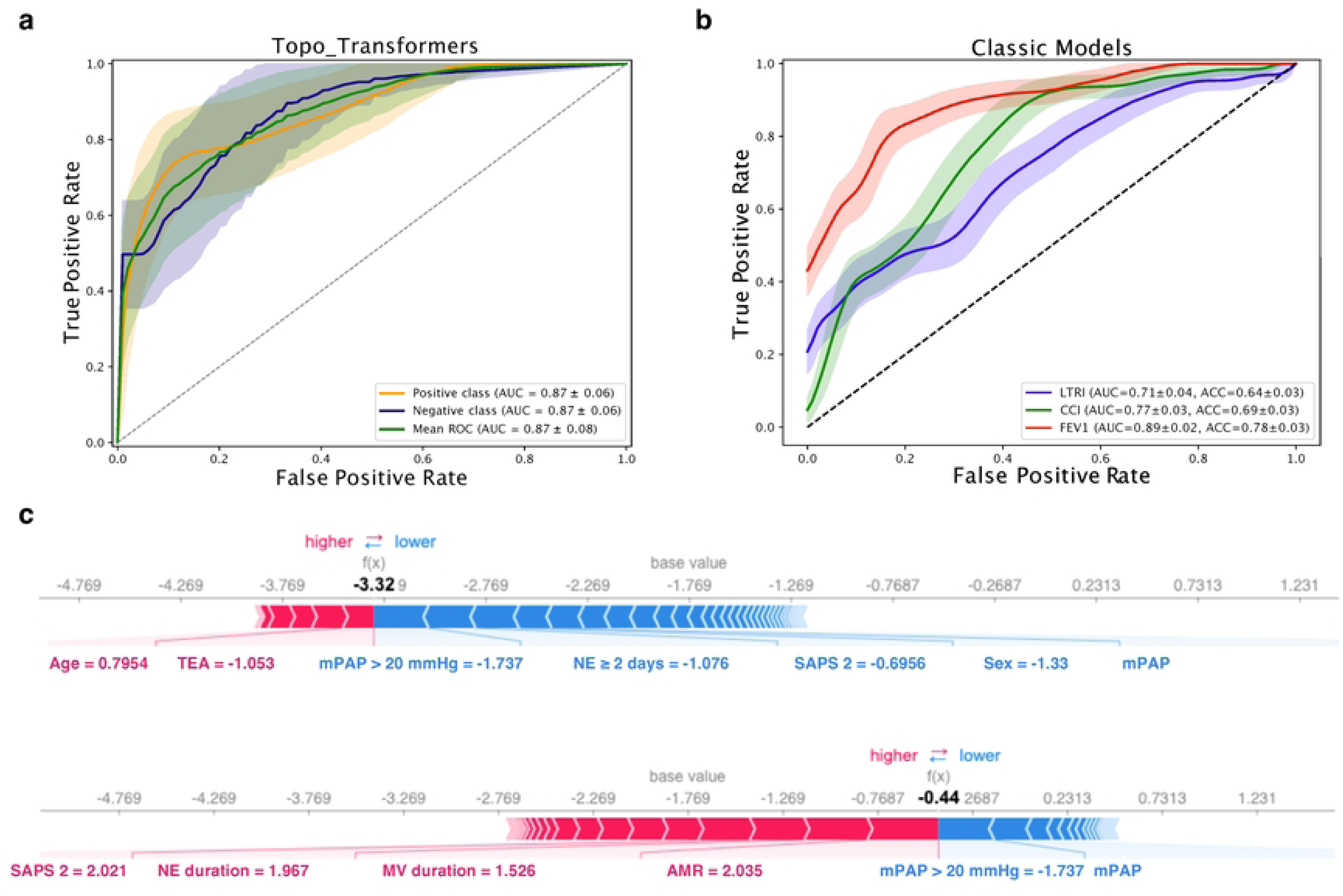
Performance comparison of the proposed method, topo transformers, with classic models in predicting the risk of mortality after lung transplantation. **a**, ROC curves with confidence intervals showing the performance of the four models. The topo transformers model outperforms the classic models with an AUC of 0.87 (std=0.06) for survivor, non-survivor classes, and mean ROC. **b**, table comparing the AUC and accuracy values for all four models. **c**, risk scores between the survivor and non-survivor groups.

To finally evaluate the performance of our score model, we compared the predicted scores to the actual outcomes for each patient. We used a variety of metrics, including accuracy, precision, recall, and F1-score, to assess the performance of the score model. We also considered any assumptions or limitations of the score model, such as potential sources of bias, and addressed them in our analysis.

Overall, the creation of this score model provided a useful tool for predicting outcomes for our patient dataset and allowed us to identify the most important features contributing to the output of our machine learning model. We make our code publicly available to ensure reproducibility.

## Results

### Cohort Characteristics and Univariate Predictors of Mortality

Our dataset, sourced from Bichat Claude Bernard’s hospital in Paris, encapsulates five years of retrospective information on 252 lung transplant patients, with a class distribution of 189 survivors and 63 non-survivors. Detailed pre-, intra-, and post-operative characteristics are presented in Table S1.

This study delves into post-transplantation mortality risk assessment through two distinct scoring frameworks. The first employs a binary risk score, simplifying outcomes into survival or impending mortality within the initial year. The second paradigm involves a nuanced temporal progression, assigning “target scores” based on mortality likelihood at 30 days, 90 days, and 1-year post-transplantation. This intricate scoring system captures the evolving mortality risk, enhancing precision.

Notably, patients aged 26-30 experienced fatal outcomes, while those under 24 consistently survived. Intensive care unit (ICU) stay duration, illustrated in Figure S1-2, reveals increased survival with time, but beyond 40 days, the risk becomes imbalanced.

Examining medical conditions, bronchial fistula emerges as a significant risk factor, with over 80% mortality. Preoperative ECMO correlates with reduced survival rates, while its use during ICU stay shows similar trends (Figure S1-2). Chronic ischemic heart disease, albeit rare, exhibits interesting patterns (Figure S3-4).

Data integrity considerations reveal 32.94% potentially containing outliers. Despite common outlier removal practices, we retained them to preserve bio-topologically vital information influencing our analysis.

### Topological Data Analysis Reveals Distinct Mortality-Associated Patterns

#### Data Lower-dimensional Visualization

Dimensionality reduction techniques (PCA, t-SNE, UMAP, PHATE) visualize high-dimensional data in lower-dimensional spaces (Fig. 2a-b). PHATE, a nonlinear technique, particularly suits high-dimensional single-cell data ^15^, applied to patient data for manifold learning (Fig. 2b). Survivor vs. non-survivor classes exhibit distinct patterns, notably in modified PHATE, contrasting with PCA, t-SNE, and UMAP.

#### Evolution of Data across Pseudo-temporal Scales

Persistence diagrams illustrate topological feature evolution across different scales ^17,18^. Survivor patients exhibit more homology generators of dimension 1, while non-survivor patients display more of dimension 2 (Fig. 2c). These features are incorporated into predictive models, enhancing outcome prediction accuracy by considering topological lung differences.

#### Evolution of Data across Spatial Scales

Transitioning from persistence diagrams to density plots (Fig. 2d) provides detailed topological feature distribution over spatial scales ^19^. Regions with larger, continuous colors indicate significant, persistent topological features. Survivor cases show fewer dimension 2 generators, suggesting a more structured topology. Metrics further support trends ^20^, indicating survivor cases’ distinct, possibly more separated clustered substructure (Fig. 3a).

#### Entropy Analysis of Persistence Diagram

Entropy analysis (Fig. 3b) assesses data variability, crucial for predicting outcomes. Near-zero concentration for dimensions 0 and 1 suggests simpler, structured topology. However, dimension 2 persistence points indicate complex, potentially significant features. Entropy analysis offers a comprehensive understanding of data complexity, contributing to precise predictive models.

#### Effect of Topological Extractors and Integration

Heatmaps (Fig. 3c-d) show the impact of topological extractors on data transformation ^21^. Comparisons reveal the significance of betti, heat kernel, persistence image, and Wasserstein metrics ^16^ for discriminating survivor and non-survivor individuals. Integration of extracted features using machine learning transformers improves classification accuracy (Fig. 4a-c). Success percentages range from 0.77 to 0.80, with red crosses indicating inadequacy. Integration enhances accuracy, employing unique 27 topological features per individual ^22^ as input for ML classification.

### Topological Transformer Model Outperforms Established Clinical Risk Scores

Figure 4c-d presents a comprehensive evaluation of different approaches, indicating successful evaluations with green or orange crosses and unsuccessful ones with red crosses. Notably, the Multi-Layer Perceptron ^22^ achieved an accuracy of 89% (see Figure 4d and Table 1), outperforming LTRI and CCI.

In Figure 4e left, the optimal learning results on patient data demonstrate a constant learning rate and momentum, achieving a perfect training set score of 1.000 and a training set loss of 0.002. While other algorithms with Nesterov’s momentum and the Adam optimizer also perform well, the inv-scaling learning rate without momentum exhibits lower scores. Calibration of the Multi-Layer Perceptron is conducted with the best parameter combination (see Figure 4d).

Figure 4e right displays an empirical cumulative distribution function (ECDF) plot of distance values between Y1 survivors and non-survivors. The survivor group consistently exhibits smaller distance values, suggesting distinctiveness between the two groups.

Figure 4f presents evaluation curves recalibrated based on the described distance. The precision-recall curve shows a sharp performance drop at a recall of 0.7 and precision of approximately 0.75, indicating potential challenges in classifying certain examples. Nevertheless, the F1 score of 0.89 reflects overall good performance. The precision-recall step function offers a detailed view, highlighting rapid precision increase until around 0.07 recall, followed by a more gradual ascent. The F1 score remains consistent at 0.89.

Finally, Figure 4g illustrates the precision-recall trade-off, showcasing a quantification of 0.83 precision and 0.80 recall. This balance suggests accurate identification of relevant cases while minimizing false positives. Despite occasional peaks, the model’s higher precision, desirable in scenarios like medical diagnoses, raises awareness of potential missed positive cases.

To our best knowledge, there are few limited models in the literature that have addressed the life-threatening issue of mortality risk after lung transplantation. Some of the existing models, such as the Lung Transplant Risk Index (LTRI), only consider preoperative variables and do not take into account any postoperative factors that may impact mortality risk. Furthermore, LTRI is only validated for patients who have undergone bilateral lung transplantation and may not accurately predict mortality risk for those who have undergone unilateral lung transplantation or other types of lung surgeries.

Other scores, such as the Forced Expiratory Volume in one second (FEV1), the Pao2/Fio2 ratio^27^, and the Charlson Comorbidity Index (CCI), have also been used to predict mortality risk after lung transplantation, but their success rates range between 64% and 78%, according to our data (see Figure 8a and Table 2). However, these scores still fall short when compared to our algorithm, which takes advantage of topological transformers and machine learning to improve its predictive performance using all available variables (see Figure 8b). Therefore, our algorithm shows promising potential as a more accurate and comprehensive tool for predicting mortality risk after lung transplantation.

**Table 2.**
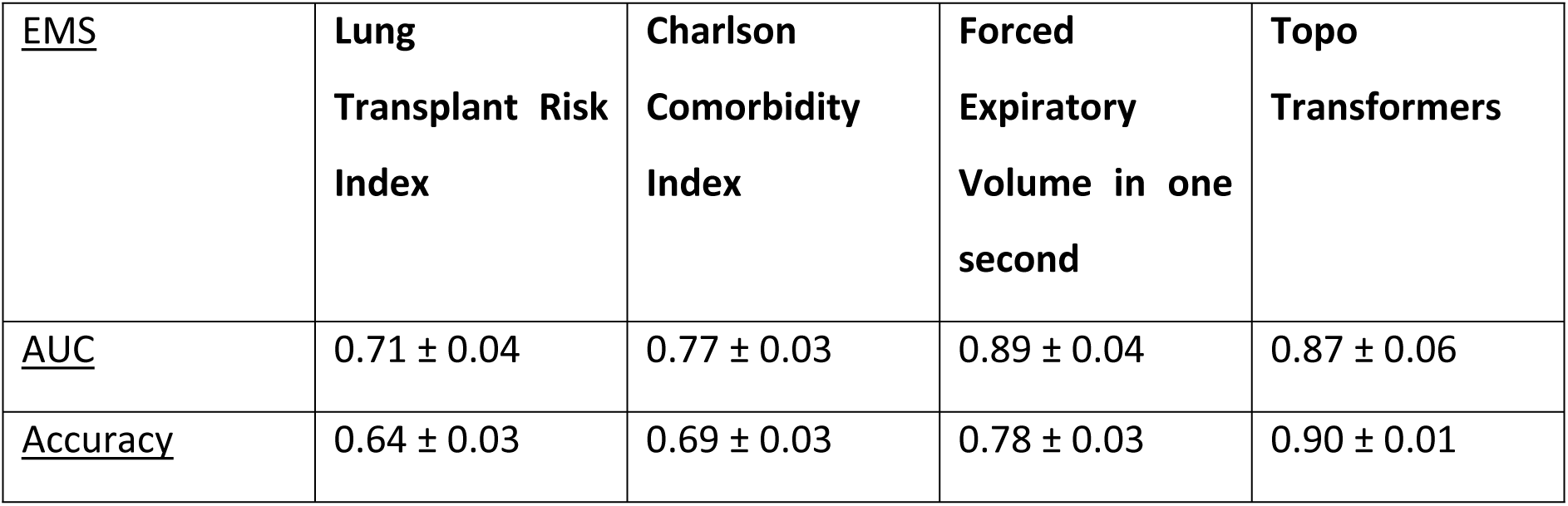
Different methods for predicting lung transplant outcomes. The methods are Lung Transplant Risk Index, Charlson Comorbidity Index, Forced Expiratory Volume in one second, and Topo Transformers. The table displays the mean Area Under the Curve (AUC) and Accuracy, with standard deviation, over 2000 bootstrap samples. AUC measures the ability of the method to distinguish between positive and negative outcomes, while Accuracy measures the proportion of correct predictions. Higher values for both AUC and Accuracy indicate better performance. It is important to note that the Pao2/Fio2 ratio was not available for analysis.

| <u>EMS</u> | **Lung**<br>**Transplant Risk**<br>**Index** | **Charlson**<br>**Comorbidity**<br>**Index** | **Forced**<br>**Expiratory**<br>**Volume in one**<br>**second** | **Topo**<br>**Transformers** |
| --- | --- | --- | --- | --- |
| <u>AUC</u> | 0.71 ± 0.04 | 0.77 ± 0.03 | 0.89 ± 0.04 | 0.87 ± 0.06 |
| <u>Accuracy</u> | 0.64 ± 0.03 | 0.69 ± 0.03 | 0.78 ± 0.03 | 0.90 ± 0.01 |

### Dynamic Post-Operative Trajectories Differentiate Survivors from Non-Survivors

This study aimed to conduct an a-priori dynamic analysis of lung transplantation trajectories, focusing on clinical outcomes such as lung function, survival, and rejection incidence. Variables of interest included pre-operative and perioperative Extracorporeal Membrane Oxygenation (ECMO), ECMO in the Intensive Care Unit (ICU), duration of ECMO use, primary graft dysfunction of grade 3, acute renal failure, and history of rejection within the past year.

ECMO therapy, an invasive treatment for severe respiratory or cardiac failure, plays a crucial role in lung transplantation outcomes. Post-operative ECMO use is associated with an increased 1-year mortality risk. Trajectory analysis revealed visually distinct behaviors in survivor and non-survivor individuals aged 15-71. Survivors exhibited stable clinical trajectories, whereas non-survivors displayed high entropy transitions. The ECMO duration and acute renal failure emerged as key predictors of mortality risk.

#### Variability Analysis between Survivor and non-Survivor Individuals

Figure 5b illustrates variability between survivor and non-survivor cohorts. Subplot 1 displays survival probability curves, indicating a higher survival probability for survivors throughout the study period. Subplot 2 shows standard deviation as a function of cluster labels, with distributions becoming sharper over time, suggesting increased variability. Subplot 3 combines information from Subplots 1 and 2, with vertical lines marking important events. Before the first line, there is low variability and a steady increase in mean. Between the lines, there is a sharp increase in standard deviation and a rapid mean growth, suggesting greater variability. After the second line, standard deviation decreases, indicating a period of greater consistency and predictability in the data, with values still increasing but at a slower pace.

### Interpretability Analysis Identifies Key Drivers of Mortality Risk

In this section, our goal is to interpret our machine learning model on a global level. We aim to identify the features that drive the model’s predictions for individual instances in the cohort, and how the impacts of these features vary across the dataset. To accomplish this, we provide a comprehensive set of model insights.

#### Feature contribution to prediction

Figure 6a presents a SHAP bar plot that interprets the impact of each feature on the model output, in terms of its contribution to the prediction (13). The features that increase the prediction (i.e., towards a higher risk of mortality) are shown in red, while those that decrease it (i.e., towards a lower risk of mortality) are shown in blue. Our MLP model shows that Acute Renal Failure feature exhibits the highest positive impact on the mortality prediction, followed closely by mechanical ventilation duration, Simplified Acute Physiologic Score 2 (SAPS2) at Day 0 (postoperative ICU admission), ICU stay duration, and antibody-mediated rejection in the past year, completing the top 5 features having the largest impact on the predictions in mean absolute value.

Complementary Figure 6b displays a heatmap of the top 10 features, individually, while grouping the remaining 39. The heatmap is arranged such that each column represents a single instance in the dataset, while each row represents a single feature. The cells in the heatmap indicate the SHAP values for each feature-instance pair. Positive values are typically shown in shades of red, while negative values are represented in shades of blue. This visualization allows for a more granular understanding of how each feature contributes to the model’s output for each individual instance in the dataset.

In our case, the instances seem to be well-separated, indicating that there may be distinct clusters of instances with similar feature values. The strong red nuance amid blue values may suggest that, for those instances, the feature represented by that column had a particularly strong positive impact on the model’s prediction, while the other features had a relatively weaker impact. These few instances can be identified with an outlier pattern in the cohort. We can use these inpatients to locally calibrate the power of our model interpretations.

The next three plots, i.e., Figure 6c-e reflect the local calibration used in our downstream analysis. In particular, Figure 6c is a SHAP force plot that describes the feature contributions to the model prediction for each individual instance in the cohort^14^. The mostly flat mountain in the middle of the plot indicates that the majority of instances have a relatively balanced mix of positive and negative feature contributions, resulting in a relatively neutral model prediction.

The winding valleys and mountains at the beginning of the force plot suggest that there is a subset of instances where the model’s prediction is driven by a complex interplay of positive and negative feature contributions, leading to more variability in the model output. This could indicate that these instances are more difficult to predict accurately and may require further investigation.

The smoother transition between valleys and mountains in the remaining 1/3 of instances at the end of the plot could suggest that these instances have more distinct and less nuanced feature contributions, resulting in a more consistent and predictable model output. Overall, the SHAP force plot provides a detailed and nuanced view of the feature contributions to the model predictions for each individual instance, which can be useful for understanding the model’s decision-making process and identifying areas for improvement.

To gain further insight into the model’s decision-making process, we utilized the dependence plots^28^ depicted in Figure 6d-e. In Figure 6d, we illustrate the local feature interactions for the top 3 most impactful variables at only one different interacting feature value each time. The last column represents the number of days on ECMO in ICU, which is an important variable proposed to be dynamically associated with a high risk of mortality after lung transplantation.

The SHAP dependence plot in Figure 6d reveals that “Acute renal failure” has major interactions with the three selected features, and its impact on the model’s prediction varies depending on the value of the feature. The flat distribution of points for most instances suggests that the feature may not be as crucial for the model’s prediction when considering these specific interactions. However, the complex cloud of points for certain feature values highlights the need for further investigation to understand the role of “Acute renal failure” in the model’s decision-making process.

It is essential to note that the feature with the largest impact on the bar plot does not necessarily have the strongest interaction with other features. The bar plot shows the absolute average contribution of each feature to the model’s output, while the SHAP dependence plot shows the interaction strength between the selected feature and other features. Therefore, both plots provide valuable insights into the model’s decision-making process, and both should be considered when interpreting the model’s behavior.

Regarding the other three features, the increasing almost linear curve with reds and blues well separated most of the time indicates that these features have a clear and consistent impact on the model’s predictions. The separation of the red and blue points suggests that the values of these features are affecting the prediction in a predictable manner. However, the occasional majority of blue or red points could indicate some outlier instances where the impact of these features on the prediction is not consistent with the general trend. Investigating these outliers further is crucial to determining their cause and whether they significantly impact the model performance. This information is used to refine the model and improve its accuracy.

Finally, Figure 6e shows a first row with a similar interpretation to those features described in the last three columns of subplot d. However, the dependence plot for ICU stay duration has an L-shape, and the color of the points is all over the place, suggesting a complex relationship between ICU stay duration and the interacting feature(s). The L-shape indicates that there is a threshold value for the interacting feature(s) beyond which the effect on ICU stay duration changes abruptly. The scatter of colors all over the place suggests that other features may interact with ICU stay duration in complex ways, causing the variation in the effect of the interacting feature(s) on ICU stay duration. Understanding the relationships between the variables in this scenario can be challenging, and further analysis may be necessary. Additionally, it’s important to note that the L-shape and scatter of colors may be influenced by the choice of the interacting feature(s) and the range of values selected for the plot.

#### Contribution to model output

Figure 7a shows the impact of different features on the model output. The red values indicate positive feature values that have a positive impact on the model output, while the remaining values are more centered around 0 and have a less significant impact.

The top 5 features (i.e., acute renal failure, mechanical ventilation duration, SAPS2 at Day 0, ICU stay duration, and antibody-mediated rejection within one year of lung transplantation) are the ones that have the highest SHAP values and are less symmetric with respect to 0. This suggests that they have a more significant impact on the model output.

However, there are some features that have positive feature values but negative SHAP values, which indicates that they have a negative impact on the model output. These features include thoracic epidural analgesia, *Cytomegalovirus* mismatch, and postoperative pneumonia.

The features SAPS2 at Day 0, ICU stay duration, mean pulmonary arterial pressure, age, and intraoperative transfusion > 2 red blood cells units have both positive and negative feature values associated with negative SHAP values, suggesting that they have a mixed impact on the model output.

To interpret Figure 7b, we must do the comparison between the swarm plot, shap bar plot, and Cohen’s distance effect bar plot^21^. One can focus on the features that appear in all three plots. These are the most important features for the model output and may provide insight into what factors are most strongly related to the predicted outcome.

Starting with the swarm plot, the top five features have the largest positive impact on the model output and are less symmetric around 0 shap value. The remaining features are more centered around 0 and have a more symmetric distribution of shap values.

Moving to the shap bar plot, you can see which features have the largest absolute shap values, indicating the features with the greatest influence on the model output. These may or may not be the same as the top features in the swarm plot, as the shap values take into account both the direction and magnitude of the feature effect.

Finally, in the Cohen’s distance effect bar plot (see Figure 7b), you can see which features have the largest effect sizes on the predicted outcome, regardless of the direction of the effect (see methods). The negative effect sizes indicate a negative relationship with the outcome, while the positive values suggest a positive relationship. Features with a value of 0 have no relationship with the outcome.

By examining all three plots, consistent important features can be identified, such as thoracic epidural Analgesia. While this feature has a positive effect on the model output in the swarm plot and a positive Cohen’s distance effect value, its shap value in the shap bar plot is negative. This suggests that thoracic epidural Analgesia is a significant predictor of the outcome, but its effect may not be straightforward and could interact with other features.

Similarly, an etiology of chronic obstructive pulmonary disease (COPD) has a positive Cohen’s distance effect value, indicating a positive relationship with the outcome, while its shap value is negative. This could indicate that the effect of an etiology of COPD on the outcome is more complex than a simple linear relationship. To explore this further, partial dependence plots were used to identify important features for the model’s performance with potential nonlinearities or interactions between features.

Figure 7c-d show how the predicted probability of an outcome changes as a feature variable changes while holding all other variables constant. The shaded regions represent the level of confidence in the model’s predictions, with a wider shaded region indicating greater variability in the model’s predictions. The subplots for acute renal failure, mechanical ventilation duration, SAPS2 at Day 0, ICU stay duration, and ECMO duration in ICU indicate that as the values of these features change, the predicted probability of mortality also changes.

Wider shaded regions may indicate that the model is less certain about its predictions for certain ranges of the feature variable. This could be due to a lack of data in that range or because the relationship between the feature and the outcome is more complex than the model can capture. Figure 7d shows the same for five more feature variables, including Age and the recently discovered important etiology of COPD variable. The subplots for some of these features, such as SAPS2 at Day 0 and ICU stay duration, suggest the presence of non-linear relationships.

However, it is important to note that the subplots only show partial dependence between a single feature and the model output, and non-linear relationships between multiple features could also be present. Overall, these findings suggest that the relationship between the model features and the outcome is complex and may require further investigation.

### Performance Comparison and Scores in Predicting Risk Mortality Y1 after Lung Transplantation

The Figure 8 showcases the performance of our proposed method, topo transformers, compared to other classic models in predicting the risk of mortality after lung transplantation. In the first subplot, the ROC curves with confidence intervals are presented, indicating that our method outperforms the three classic models, with an AUC of 0.87 (std=0.06) for survivor, non-survivor classes, and mean ROC. The AUC values for the three classic models were 0.71 (std=0.04) for LTRI, 0.77 (std=0.03) for CCI, and 0.89 (std=0.02) for FEV1.

The second subplot presents a table comparing the AUC and accuracy values for all four models, including our proposed method and the three classic models. The topo transformers model achieves an AUC of 0.87 (std=0.06) and an accuracy of 0.90 (std=0.01), which are the highest values among all models. This table clearly shows that our proposed method outperforms the classic models in terms of both AUC and accuracy.

Finally, the third plot shows a comparison of the risk scores between the survivor and non-survivor groups, as visualized in a SHAP force plot. This plot is based on all the information previously discussed and shows the contribution of each feature to the prediction of the risk of mortality after lung transplantation. The risk scores are higher for the non-survivor group compared to the survivor group, indicating that the features have a stronger impact on the risk of mortality for the non-survivor group.

## Discussion

The findings of our study demonstrate the potential of topological transformers and ML in enhancing mortality risk prediction after lung transplantation. By integrating both preoperative and postoperative variables, our algorithm achieves superior predictive accuracy (up to 90%) compared to traditional models such as the Lung Transplant Risk Index (LTRI) and the Charlson Comorbidity Index (CCI). This improvement underscores the limitations of existing risk scores^29^, which rely heavily on static preoperative factors and fail to account for dynamic postoperative changes that significantly influence patient outcomes.

### Comparison with Existing Literature and Clinical Relevance

Previous studies have primarily focused on preoperative risk stratification in lung transplantation. The LTRI, for instance, incorporates variables such as age, diagnosis, and pulmonary artery pressure but lacks postoperative data, limiting its real-world applicability^30^. Similarly, the CCI, a widely used comorbidity index, does not account for transplant-specific complications or evolving clinical trajectories^31^. Recent advances in ML have enabled more dynamic risk assessment, with studies demonstrating improved predictive performance when incorporating postoperative variables^32^. Our work aligns with these findings but extends them by leveraging TDA to uncover non-linear relationships between clinical features and outcomes^33^, a novel contribution to the field.

### Methodological Innovations and the Role of Topological Data

A key strength of our model is its ability to adapt to changes in a patient’s clinical course. Postoperative variables, such as the need for extracorporeal membrane oxygenation (ECMO), mechanical ventilation, or vasopressor support, were among the top predictors of mortality, as indicated by SHAP analysis. While some of these factors (e.g., ECMO use) reflect disease severity and are non-modifiable, others (e.g., antibody-mediated rejection or pain management strategies) present opportunities for intervention. For example, earlier detection of rejection episodes or optimized analgesia (e.g., thoracic epidural) could mitigate complications, underscoring the clinical utility of real-time risk monitoring.

This dynamic approach aligns with emerging trends in precision medicine, where continuous data integration allows for personalized treatment adjustments^34^. Similar ML-based dynamic models have shown promise in other organ transplants, such as liver and heart transplantation^35^, suggesting broad applicability beyond lung transplantation.

### Limitations and Future Directions

Our use of topological transformers represents a methodological advance in handling complex medical data. Unlike conventional ML models that may overlook intricate interactions between variables, TDA captures high-dimensional patterns, improving feature extraction and interpretability^16^. The partial dependence plots in our study revealed non-linear associations between certain predictors and mortality risk, reinforcing the need for advanced modeling techniques in clinical prediction.

However, our study has limitations, including its single-center, retrospective design. Multicenter validation is essential to ensure generalizability, particularly given variations in post-transplant care protocols. Additionally, ethical considerations—such as algorithmic bias, transparency, and integration into clinical workflows—must be addressed before widespread adoption^36^. Future research should explore real-time implementation, possibly through digital health platforms that continuously update risk predictions based on evolving patient data.

## Conclusion

This study introduces a topological transformer-based predictive model for post-transplantation mortality. By incorporating dynamic clinical features and leveraging advanced ML techniques, the model achieves superior accuracy compared to existing predictive tools. Future work will focus on real-time implementation and broader clinical validation.

## Research ethics approval

This work received approval from the Ethics Committee at the Bitchat Hospital, Université de Paris Cité.

## Ethics Statement

This study was approved by the Partners Human Research Committee, which waived the requirement for informed consent for the collection and analysis of samples.

## CRediT Authorship Contribution Statement

A Tran-Dinh and E Atchade performed data curation and formal analysis. A Tran-Dinh contributed to the methodology, writing of the original draft, and review and editing of the manuscript. I Morilla contributed to the conceptualization, data curation, formal analysis, supervision, funding acquisition, investigation, visualization, methodology, project administration, and writing of the original draft, as well as review and editing of the manuscript with contribution of all authors.

## Data Availability

All data used in this work are included in the article and/or supplementary material.

## Code availability

The code for the implementation of the proposed method is available on GitHub at https://github.com/MorillaLab/TopoTransformers/Models. Additionally, the code used for the analysis and experiments presented in this paper is also available on GitHub at https://github.com/MorillaLab/TopoTransformers/Analysis.

## Supplementary Information

Additional information is available in the supplementary materials, which can be found at repository.

## Acknowledgements

We gratefully acknowledge funding from the National Research Association (ANR) (Inflamex renewal 10-LABX-0017 to I Morilla), Consejería de Universidades, Ciencias y Desarrollo, fondos FEDER de la Junta de Andalucía (ProyExec_0499 to I Morilla), DHU FIRE Emergence 4, and l’Agence de la Biomedecine (to A Tran-Dinh).

## Abbreviations

ACR: Acute cellular rejection within one year of LT
AMR: Antibody-mediated rejection within one year of LT
BAD: Bronchial anastomosis dehiscence within one year of LT
BMI: Body mass index
CAD: Coronary artery disease
CMV Mismatch: *Cytomegalovirus* mismatch (Recipient −/Donor +)
COPD: Chronic obstructive pulmonary disease
ECMO: Extracorporeal membrane oxygenation Grade 3
PGD: grade 3 primary graft dysfunction
HELT: High emergency lung transplantation
ICU_LOS: Intensive care unit length of stay
Intraoperative Transfusion: Intraoperative transfusion > 2 red blood cells units
LT procedure: LT : lung transplantation, 0 = single LT, 1= double LT
mPAP: Mean pulmonary arterial pressure
NE: Norepinephrine
NE duration: Norepinephrine duration
PE: Pleural empyema within one year of LT
Pneumonia_ESBL: pneumonia caused by extended-spectrum beta-lactamase-producing bacteria
RRT: Renal replacement therapy in ICU
SAPS 2: Simplified Acute Physiology Score 2 at postoperative ICU admission
Sex: 0 = female, 1 = male
SOFA_score_Day_0: SOFA score at postoperative ICU admission
TEA: Thoracic epidural analgesia
VAP ESBL PA: Ventilation acquired pneumonia du to ESBL producing bacteria or Pseudomonas aeruginosa

## Supplemental figures legends

Figure S1. **Distribution Analysis of Patients Based on Binary Risk Mortality.** Histogram distributions of cohort variables are presented in each panel, illustrating the statistical analysis of patients categorized according to binary risk mortality.

Figure S2. **Distribution Analysis of Patients Based on “Target Scores” Risk Mortality.** Histogram distributions of different variables are displayed in each panel, elucidating the statistical analysis of patients categorized based on “target scores” risk mortality.

Figure S3. **Boxplot Analysis of Patients Based on Binary Risk Mortality.** Boxplot distributions of respective variables are exhibited in each panel, providing insights into the statistical analysis of patients categorized by binary risk mortality.

Figure S4. **Boxplot Analysis of Patients Based on “Target Scores” Risk Mortality.** Boxplot distributions of corresponding variables are showcased in each panel, shedding light on the statistical analysis of patients categorized by “target scores” risk mortality.

Table S1. **Patient Cohort Description.** This table offers a comprehensive description of the patient cohort. It encompasses individual donor factors, transplant procedural and recipient factors, post-transplant complications, and the innovative TDA-ML approach employed for predicting mortality risk.

